# Study partner profile effects on CDR-SB change in anti-amyloid therapy evaluation

**DOI:** 10.64898/2026.06.22.26356066

**Authors:** Ambre Mounié, Kenichiro Sato, Saki Nakashima, Yoshiki Niimi, Takeshi Iwatsubo

**Affiliations:** Dementia Inclusion and Therapeutics, The University of Tokyo Hospital, Tokyo, Japan; Cognitive Sciences, Université Côte d’Azur, Nice, France; Unit for Early and Exploratory Clinical Development, The University of Tokyo Hospital, Tokyo, Japan; Department of Neurology, The University of Tokyo Hospital, Tokyo, Japan; National Center of Neurology and Psychiatry, Tokyo, Japan

**Keywords:** Alzheimer’s disease, Clinical Dementia Rating Sum of Boxes, study partner, informant, anti-amyloid therapy, bias analysis

## Abstract

**INTRODUCTION:** The Clinical Dementia Rating Sum of Boxes (CDR-SB), a primary outcome in anti-amyloid therapy (AAT) trials, integrates information from participants and study partners. CDR-SB scores may vary by study partner characteristics, but their impact on 18-month change interpretation remains unclear.

**METHODS:** Using the NACC Uniform Data Set, we fitted linear mixed-effects calibration models in an Alzheimer’s disease (AD)-primary early symptomatic cohort and propagated study partner-associated coefficients through Monte Carlo simulations. We estimated components of 18-month CDR-SB change under observed profile changes, simulated follow-up imbalance in a common female living-with profile, and tipping-point scenarios. Analyses were repeated in amyloid-positive and trial-like cohorts.

**RESULTS:** The AD-primary cohort included 15,061 participants and 7,683 baseline-to-18-month pairs. Observed profile changes generated a negligible cohort-level component (mean 0.0014 points, 95% simulation interval 0.0006 to 0.0022). Simulated follow-up imbalance generated differences of 0.014 to 0.071 points across 10% to 50% reassignment. Under the primary calibration model, generating a 0.45-point difference, equal to the reported Clarity AD CDR-SB group difference, required median net imbalance >100% and was feasible in 48% of iterations. Amyloid-positive and trial-like cohorts had lower median tipping points but wider intervals, reflecting coefficient imprecision.

**DISCUSSION:** In the large AD-primary cohort, observed study partner profile changes and simulated follow-up imbalance generated CDR-SB differences that were small relative to the 0.45-point Clarity AD benchmark. Biomarker-confirmed estimates were less stable because of coefficient imprecision. These findings suggest limited impact under typical AD-primary conditions but support systematic study partner profile collection and sensitivity analyses in observational and external-comparator CDR-SB studies for AAT evaluation.

## Introduction

The approval of anti-amyloid therapies (AAT) for early Alzheimer’s disease (AD) has marked a turning point in the clinical management of the disease (1). In the pivotal Clarity AD trial, lecanemab demonstrated a statistically significant slowing of clinical decline, with an adjusted mean difference of -0.45 points on the Clinical Dementia Rating Sum of Boxes (CDR-SB) at 18 months compared to placebo (2). Donanemab also slowed clinical decline in early symptomatic AD, although the primary endpoint in TRAILBLAZER-ALZ 2 was the integrated Alzheimer’s Disease Rating Scale, with CDR-SB assessed as a key secondary outcome (3). Together with prior work supporting CDR-SB as a trial endpoint (4), these findings show the importance of CDR-SB interpretation in AAT evaluation.

The CDR-SB is a structured composite measure rated across six cognitive and functional domains: memory, orientation, judgment and problem solving, community affairs, home and hobbies, and personal care, each rated on a five-point scale (5). Its psychometric properties make it well-suited as a primary endpoint, and it allows for smaller sample sizes compared to alternative measures (4,6). However, the CDR-SB is derived from a structured clinical interview conducted with both the patient and an informant (4,5). This structural dependence on an external informant introduces a source of variability and bias that is not related to the patient’s actual disease severity (7).

A growing body of evidence has begun to quantify how informant characteristics systematically influence CDR-SB scores. After adjustment for patient clinical severity, female informant sex, adult child relationship, daily contact, and cohabitation have each been associated with higher CDR-SB scores (8). Beyond cross-sectional associations, informant replacement in AD clinical trials has been associated with bias toward worse performance in functional outcomes, even when it did not significantly affect mean change from baseline (7). Study partner type has also been linked to differences in observed CDR-SB progression rates (9). However, it remains unclear how much 18-month CDR-SB change could be influenced by broader study partner profile attributes, baseline-to-follow-up profile changes, or between-arm profile imbalance.

Several practical questions therefore remain unanswered. It is unclear whether naturally occurring baseline-to-follow-up study partner profile changes meaningfully affect 18-month CDR-SB change estimates, whether differential follow-up imbalance could create clinically relevant between-group differences, and what degree of imbalance would be required to approach the reported lecanemab treatment effect. These questions are particularly important for real-world registries, external-comparator studies, and target trial emulations of AAT, where randomization cannot be relied upon to balance study partner profiles (10).

To address these questions, we used the NACC Uniform Data Set to estimate associations between study partner profile characteristics and CDR-SB levels in a strict AD-primary early symptomatic cohort, building on prior NACC work (8). We then propagated these calibration associations, which do not represent causal effects of changing study partners, through Monte Carlo simulations to estimate study partner-associated components of 18-month CDR-SB change under observed profile changes and simulated follow-up imbalance scenarios. Analyses were repeated in amyloid-positive and trial-like subsets relevant to anti-amyloid therapy evaluation.

## Methods

### Study design and participants

This study used the National Alzheimer’s Coordinating Center (NACC) Uniform Data Set (UDS), a multicenter longitudinal registry comprising data collected across Alzheimer’s Disease Research Centers in the United States (11). The analytic dataset included UDS form versions 1.2 through 4. All visits available for eligible participants were included in the calibration model, and baseline-to-follow-up pairs were constructed for the simulation analyses as described below: The primary analytic cohort comprised participants with MCI or mild dementia at baseline, defined using *NACCUDSD* values of 3 or 4, global CDR values of 0.5 or 1 (*CDRGLOB*), CDR-SB values between 0.5 and 9 (*CDRSUM*), and AD as the primary etiologic diagnosis, defined as *NACCETPR* = 1 or *NACCALZP* = 1. This cohort included participants with unknown or negative amyloid biomarker status, as the primary inclusion criterion was clinical diagnosis rather than biomarker confirmation.

Three secondary cohorts reflected populations more directly relevant to the anti-amyloid treatment era. The amyloid-positive cohort included participants from the primary cohort with confirmed amyloid positivity, defined as a positive amyloid PET (*AMYLPET* = 1) or CSF result (*AMYLCSF* = 1). Two trial-like cohorts, nested within the amyloid-positive cohort, additionally applied MMSE-equivalent eligibility score restrictions of 20–30 and 22–30, respectively. These ranges approximate commonly used cognitive eligibility ranges in AAT trial-eligibility studies rather than exactly reproducing individual trial criteria (12,13). These restrictions used an MMSE-equivalent eligibility variable, constructed from available MMSE and incorporating MoCA-to-MMSE conversion when MMSE was unavailable. Because amyloid biomarker availability in NACC is selective and reflects center-level practices, these cohorts are analyzed as secondary rather than primary estimation sets.

The availability of key variables including the primary outcome CDR-SB (score ranging from 0 to 18), informant sex, relationship, contact frequency, and cognitive scores was assessed for each cohort before model fitting. Calibration models were fitted using complete-case visits for CDR-SB, zCog, informant sex, relationship, contact frequency, participant sex, visit time, participant identifier, and center identifier.

### Cognitive severity covariate

A composite cognitive severity variable, zCog, was constructed for use as a covariate in the calibration model. Raw MMSE and raw MoCA scores were standardized separately using the mean and standard deviation computed within the AD-primary calibration cohort. When both scores were available at a given visit, the MMSE z-score was used; when MMSE was missing, the MoCA z-score was used. This standardization was anchored to the primary cohort to ensure comparability across secondary cohorts.

### Study partner profile variables

NACC UDS uses the term co-participant, whereas the clinical-trial literature often uses study partner; we use study partner as the main term and retain informant when referring to prior literature or original variable labels. Three study partner profile dimensions were coded for each visit. Informant sex was coded as a binary variable: female (1) versus male (0), derived from *INSEX*. Relationship to the participant was coded as a three-category variable derived from *INRELTO*: spouse or partner (reference category), adult child, and other.

Contact frequency and cohabitation were coded as a four-category variable using a source-aware hierarchical approach. The four categories were: less than weekly (reference), weekly or more, daily, and lives-with (see Supplementary Methods). The reference profile for the simulation analyses was defined as: male informant, spouse or partner relationship, less-than-weekly contact. This profile served as a modeling reference category for defining profile contrasts, not as a representative real-world study partner profile.

### Follow-up selection

For each eligible participant, the baseline visit was defined as the first chronological visit meeting all inclusion criteria. The follow-up visit was defined as the visit closest to 18 months within a window of 12 to 24 months from baseline. When multiple visits were equidistant from the 18-month target, the visit occurring before 18 months was preferred. Only participants with complete data on CDR-SB, informant sex, relationship, and contact frequency at both baseline and follow-up were retained for the simulation analyses. Visits with missing contact frequency information were excluded from both the calibration model and the simulation analyses.

### Calibration model

Following the general approach of Vargas-Gonzalez et al. (8), we fitted a linear mixed-effects calibration model to all visits of participants in the primary AD-primary cohort to estimate associations between study partner profile characteristics and CDR-SB levels. The model specification was: *CDR-SB ∼ time_years + zCog + pt_female + inf_female + rel_3cat + contact_4cat + pt_female:rel_3cat + (1 + time_years || NACCID) + (1 | NACCADC)*. Fixed effects included time since first visit in years, a standardized cognitive severity score, participant sex, informant sex, relationship category (adult child and other, each relative to spouse or partner), contact frequency category (weekly-or-more, daily, and lives-with, each relative to less than weekly), and the interaction between participant sex and relationship. Random effects included a participant-level random intercept and random slope for time, and a center-level random intercept.

The model was fitted using restricted maximum likelihood via the *lme4* package in R, with the *bobyqa* optimizer, and was fitted separately in each AAT-relevant secondary cohort. Details of the contact frequency coding hierarchy and the model convergence fallback sequence are provided in Supplementary Methods.

The coefficients from the calibration model are calibration associations, not causal effects of changing a study partner. We therefore interpret simulation outputs as study partner-associated CDR-SB components, not as measurement bias estimates or causal switching effects.

### Monte Carlo sensitivity analysis

The study partner-associated coefficient vector β and its variance-covariance matrix V were extracted from the calibration model. At each Monte Carlo iteration, a coefficient vector β(m) was drawn from a multivariate normal distribution with mean β and covariance matrix V. For each participant, the study partner-associated component of 18-month CDR-SB change was computed as: *component_i(m) =* Δ*X_i ·* β*(m)*, where ΔX_i represents the baseline-to-follow-up change in study partner profile. Scenarios A through D used 10,000 Monte Carlo iterations for each cohort. The supplementary uncertainty-decomposition analysis used 2,000 iterations per beta-sampling condition and pseudo-arm size.

### Simulation scenarios

Scenario A, Observed NACC profile changes: The observed design vector differences ΔX derived from actual baseline-to-follow-up study partner profile changes in NACC were used directly. This scenario estimated the study partner-associated component of 18-month CDR-SB change under naturally occurring profile changes.

Scenario B, Simulated follow-up imbalance in the female living-with profile: Two pseudo-arms of n=250 participants each were constructed by bootstrap sampling with replacement from the same pool of baseline-to-follow-up pairs. This pseudo-arm size was chosen as a pragmatic external-comparator sample size; the influence of pseudo-arm size on simulation interval width was evaluated in Table S2D. In the imbalanced comparator arm, 10%, 20%, 30%, or 50% of participants were reassigned at follow-up to the female living-with target profile, with 5% results provided in the Supplementary Materials. The study partner-associated CDR-SB difference was computed as the mean component in the imbalanced comparator arm minus the reference arm. The female living-with profile was selected because it was common in the analytic dataset, observed in approximately 47% of visits. Baseline profiles were unchanged; reassignment was applied only at follow-up.

Scenario C, Female adult-child living-with stress-test enrichment: The same procedure was applied to the prespecified female adult-child living-with profile. Because this complete profile was observed in only approximately 4% of participants, we interpreted this scenario as an upper-bound stress test rather than as a plausible observational imbalance.

Scenario D, Tipping-point analysis: For each prespecified CDR-SB difference threshold (0.10, 0.20, 0.30, and 0.45 points), we estimated the net study partner profile imbalance required to generate a study partner-associated difference of that magnitude. The main tipping-point analysis used the female living-with contrast, comparing the reference profile (male informant, spouse or partner, less than weekly contact) to the target profile (female informant, spouse or partner, lives-with contact). A supplementary maximal tipping-point analysis used the female adult-child living-with profile contrast. Results are reported as the median required net imbalance and the 2.5th to 97.5th percentile simulation interval across Monte Carlo iterations.

The decomposition of simulation uncertainty was applied to both the primary cohort and all AAT-relevant secondary cohorts, see details in Supplementary Method.

### Sensitivity analyses

Prespecified sensitivity analyses including an expanded calibration model, alternative follow-up window, quadratic cognitive severity term, exclusion of unreliable informant visits, participant fixed-effect model, change-on-change model, and external coefficient sensitivity analysis are described in Supplementary Methods.

All analyses were conducted in R version 4.5.1 (2025-06-13). Mixed-effects models were fitted using the lme4 (14) and lmerTest packages. Monte Carlo sampling was performed using the MASS package. Session information was recorded at the start and end of each analysis run. Random seeds were anchored at 260530 with cohort- and scenario-specific offsets to ensure reproducibility across analyses.

### Ethics

This study was approved by the University of Tokyo Graduate School of Medicine institutional ethics committee (ID: 2025264NI). Informed consent was not required because the study uses anonymized, publicly-available data only.

## Results

### Cohort characteristics

The primary AD-primary cohort included 15,061 participants across 46 centers, contributing 47,958 visits, of whom 7,683 had complete baseline-to-18-month follow-up pairs available for simulation analyses. The amyloid-positive cohort included 1,505 participants (36 centers, 3,858 visits, 728 pairs), the Aβ+ MMSE-equivalent 20-30 cohort included 1,190 participants (588 pairs), and the Aβ+ MMSE-equivalent 22-30 cohort included 1,042 participants (518 pairs).

The female living-with profile was common across cohorts, whereas the female adult-child living-with stress-test profile was rare. Informant replacement was infrequent, and profile attribute changes were modest. Cohort characteristics and study partner profile prevalence are summarized in Table 1.

**Table 1:** Cohort characteristics and observed study partner profile prevalence. Demographic and baseline clinical characteristics are summarized at the participant level using baseline visits. Study partner profile variables are summarized at the visit level across all available visits within each cohort. Percentages for study partner profile variables are computed at the visit level across all available visits within each cohort. Baseline-to-18-month pairs represent participants with complete data on CDR-SB, informant sex, relationship, and contact frequency at both baseline and at the follow-up visit closest to 18 months within a 12-to-24-month window. Profile prevalence percentages reflect the pooled baseline and follow-up distribution. Any profile attribute change indicates that at least one of informant sex, relationship, or contact frequency differed between baseline and follow-up. Informant replacement at follow-up is based on the NEWINF variable. The standardized cognitive score (zCog) was constructed from raw MMSE and raw MoCA scores standardized separately within the AD-primary calibration cohort. The MMSE-equivalent eligibility score was used exclusively for defining trial-like AAT eligibility cuts and may incorporate MoCA-to-MMSE conversion. The AD-primary cohort was substantially larger than the AAT-relevant secondary cohorts, providing more stable coefficient estimates. The female living-with profile was common across all cohorts (47–50% of visits), supporting its use as the main simulated between-arm imbalance scenario in observational settings. In contrast, the female adult-child living-with profile remained rare (2–4% of visits), supporting its interpretation as an upper-bound stress-test scenario rather than a plausible distributional difference. **Abbreviations:** AD, Alzheimer’s disease; Aβ+, amyloid-positive; MMSE-eq, MMSE-equivalent eligibility score range used for cohort definition; MMSE, Mini-Mental State Examination; MoCA, Montreal Cognitive Assessment; CDR-SB, Clinical Dementia Rating Sum of Boxes; NEWINF, new informant indicator; zCog, standardized cognitive severity score; AAT, antiamyloid therapy; n, number.

| Cohort | AD-primary | Aβ+ | Aβ+ MMSE-equivalent 20-30 | Aβ+ MMSE-equivalent 22-30 |
| --- | --- | --- | --- | --- |
| Sample size, n | 15061 | 1505 | 1190 | 1042 |
| Visits, n | 47958 | 3858 | 3148 | 2784 |
| Centers, n | 46 | 36 | 36 | 36 |
| Baseline-to-18-month pairs, n | 7683 | 728 | 588 | 518 |
| Age at baseline, mean (SD) | 73.4 (9.3) | 69.0 (8.8) | 69.9 (8.5) | 70.3 (8.3) |
| Female participant, % | 53.17 | 51.10 | 50.00 | 49.33 |
| Education, years, mean (SD) | 14.9 (3.6) | 16.0 (3.0) | 16.1 (2.9) | 16.2 (3.0) |
| White or non-Hispanic White, % | 74.95 | 78.87 | 78.24 | 78.31 |
| Black or non-Hispanic Black, % | 11.92 | 4.39 | 4.54 | 4.22 |
| Hispanic, % | 8.90 | 11.89 | 12.27 | 12.48 |
| Asian, % | 2.44 | 1.33 | 1.18 | 1.25 |
| Other or missing race/ethnicity, % | 1.78 | 3.52 | 3.78 | 3.74 |
| Baseline CDR-SB, mean (SD) | 3.53 (2.16) | 3.24 (1.96) | 2.87 (1.78) | 2.66 (1.69) |
| Baseline MMSE-equivalent score, mean (SD) | 23.8 (4.2) | 24.9 (3.0) | 25.2 (2.8) | 25.9 (2.2) |
| MMSE-equivalent score availability, % | 77.28 | 64.88 | 76.14 | 78.84 |
| zCog availability, % | 81.21 | 76.98 | 82.24 | 83.73 |
| Female informant, % | 67.88 | 60.92 | 62.33 | 63.42 |
| Adult child informant, % | 22.15 | 11.19 | 10.88 | 11.49 |
| Lives-with % | 72.96 | 82.21 | 82.31 | 81.85 |
| Female living-with profile, % | 46.99 | 47.46 | 48.98 | 49.61 |
| Female adult child living-with profile, % | 4.13 | 2.27 | 2.12 | 2.22 |
| Any profile attribute change % | 15.71 | 12.36 | 12.24 | 12.55 |
| Informant replacement % | 7.08 | 4.26 | 5.1 | 5.21 |

### Calibration model coefficients

In the AD-primary cohort, all informant-associated terms except the participant-sex-by-relationship interaction were statistically distinguishable from zero. A female informant was associated with a 0.143-point higher CDR-SB score (95% CI 0.055 to 0.230), and lives-with cohabitation with a 0.303-point higher score (95% CI 0.122 to 0.483). Adult child relationship and intermediate contact frequency categories showed positive associations, smaller than the lives-with coefficient. The other-relationship category was associated with lower CDR-SB levels than spouse or partner informants in the AD-primary cohort (−0.295 points, 95% CI −0.460 to −0.131), although this term was not used as a target profile in the main simulation scenarios. Full coefficients are reported in Table S1A.

In the AAT-relevant secondary cohorts, female informant and lives-with coefficients were generally positive but substantially less precise. Relationship coefficients were not directionally consistent, with adult-child coefficients estimated below zero in the amyloid-positive subsets, reflecting instability in these smaller cohorts (Figure 1). Prespecified sensitivity analyses were broadly consistent with the main interpretation, although participant fixed-effect and change-on-change models showed less stable contact-frequency estimates (Table S1B).

**Figure 1:**
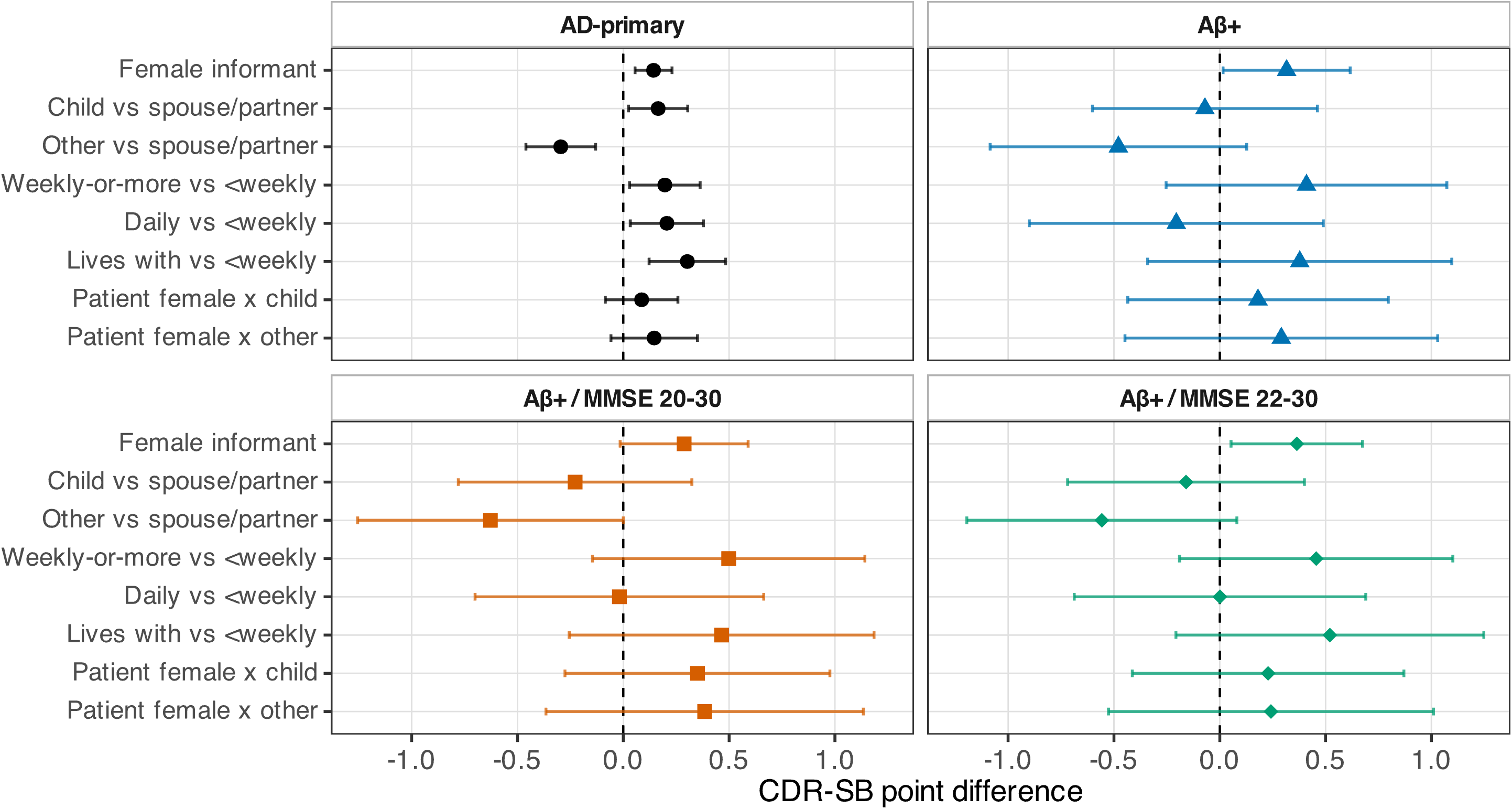
Calibration model coefficients for the association between study partner profile characteristics and CDR-SB levels, by cohort. Point estimates and approximate Wald 95% confidence intervals for study partner-associated fixed-effect terms from the primary linear mixed-effects calibration model, shown separately for each analytic cohort. A value of zero indicates no association with CDR-SB level relative to the reference category. The reference categories are male informant, spouse or partner relationship, and less-than-weekly contact. Wider confidence intervals in the amyloid-positive and trial-like cohorts reflect reduced precision due to smaller sample sizes. **Abbreviations:** CDR-SB, Clinical Dementia Rating Sum of Boxes; Aβ+, amyloid-positive; MMSE-eq, MMSE-equivalent eligibility score range used for cohort definition.

### Observed natural transitions

Naturally occurring study partner profile changes generated a negligible cohort-level study partner-associated component of 18-month CDR-SB change in the AD-primary cohort (mean 0.0014 points, 95% SI 0.0006 to 0.0022). Components in AAT-relevant cohorts were also near zero, with simulation intervals crossing or approaching zero (Table 2A).

**Table 2:** Study partner-associated CDR-SB components under observed profile changes, simulated follow-up imbalance, and tipping-point scenarios. **Table 2A.** Values in the observed-transition row represent the mean study partner-associated CDR-SB component under naturally occurring baseline-to-follow-up profile changes. Values in the reassignment rows represent the mean study partner-associated CDR-SB difference between the imbalanced comparator arm and the reference arm after follow-up reassignment to the female living-with target profile. The difference was defined as imbalanced comparator arm minus reference arm; reassignment in the reference arm instead would reverse the sign without changing the magnitude. Achieved target-profile prevalence after reassignment is reported in Table S2A. The achieved increase in female living-with prevalence in the comparator arm relative to the reference arm was equal to the requested reassignment level at 10%, 20%, and 30% across all cohorts. At 50% requested reassignment, the achieved increase was 49.7, 49.5, 49.1, and 48.9 percentage points in the AD-primary, Aβ+, Aβ+ MMSE-equivalent 20–30, and Aβ+ MMSE-equivalent 22–30 cohorts respectively. **Table 2B.** Values represent the median required net target-profile imbalance (95% simulation interval) needed to generate the specified study partner-associated CDR-SB difference. Values exceeding 100% are theoretically unachievable and indicate that the specified difference cannot be generated through this profile contrast alone under the estimated calibration coefficients. The proportion of iterations where the threshold is achievable reports the fraction of Monte Carlo iterations in which the required net imbalance was 100% or less. Values below 1.00 indicate that the specified study partner-associated difference cannot be generated in all plausible coefficient scenarios, even under complete follow-up reassignment of the comparator arm. The proportion of Monte Carlo iterations in which the required net imbalance was ≤100% was as follows. AD-primary: 1.00, 0.99, 0.92, and 0.48 at the 0.10, 0.20, 0.30, and 0.45-point thresholds respectively. Aβ+: 0.96, 0.91, 0.85, and 0.73. Aβ+ MMSE-equivalent 20–30: 0.96, 0.92, 0.87, and 0.78. Aβ+ MMSE-equivalent 22–30: 0.98, 0.96, 0.93, and 0.86. Values below 1.00 indicate that the specified study partner-associated difference cannot be generated in all plausible coefficient scenarios, even under complete follow-up reassignment of the comparator arm. **Abbreviations:** CDR-SB, Clinical Dementia Rating Sum of Boxes; Aβ+, amyloid-positive; MMSE-eq, MMSE-equivalent eligibility score range; SI, simulation interval; pp, percentage points.

| Cohort | AD-primary | Aβ+ | Aβ+ MMSE-equivalent 20-30 | Aβ+ MMSE-equivalent 22-30 |
| --- | --- | --- | --- | --- |
| Observed transition | 0.0014 (0.0006 to 0.0022) | 0.0014 (-0.0054 to 0.0081) | -0.0046 (-0.0130 to 0.0038) | -0.005 (-0.0147 to 0.0049) |
| 10% reassignment | 0.014 (-0.002 to 0.031) | 0.033 (-0.005 to 0.073) | 0.030 (-0.008 to 0.070) | 0.038 (-0.000 to 0.079) |
| 20% reassignment | 0.029 (0.008 to 0.050) | 0.066 (0.008 to 0.126) | 0.059 (0.001 to 0.119) | 0.076 (0.017 to 0.136) |
| 30% reassignment | 0.043 (0.017 to 0.069) | 0.099 (0.019 to 0.180) | 0.089 (0.010 to 0.173) | 0.114 (0.033 to 0.197) |
| 50% reassignment | 0.071 (0.034 to 0.110) | 0.164 (0.039 to 0.292) | 0.148 (0.019 to 0.278) | 0.184 (0.058 to 0.315) |

| Cohort | AD-primary | Aβ+ | Aβ+ / MMSE-eq 20-30 | Aβ+ / MMSE-eq 22-30 |
| --- | --- | --- | --- | --- |
| 0.10-point threshold | 22.5% (15.4 to 41.1%) | 14.5% (6.8 to 177.9%) | 13.4% (6.6 to 155.2%) | 11.3% (5.9 to 77.7%) |
| 0.20-point threshold | 45.0% (30.8 to 82.3%) | 29.0% (13.6 to 355.7%) | 26.7% (13.3 to 310.4%) | 22.6% (11.8 to 155.3%) |
| 0.30-point threshold | 67.4% (46.3 to 123.4%) | 43.5% (20.5 to 533.6%) | 40.1% (19.9 to 465.5%) | 34.0% (17.7 to 233.0%) |
| 0.45-point threshold | 101.1% (69.4 to 185.1%) | 65.2% (30.7 to 800.4%) | 60.1% (29.9 to 698.3%) | 50.9% (26.5 to 349.5%) |

Specific profile transitions were uniformly rare, including transitions to the female living-with profile and to the female adult-child living-with stress-test profile (Figure S1). Results were consistent using the alternative 9–27 month follow-up window and published external coefficients from the prior NACC study (8).

### Simulated follow-up imbalance in the female living-with profile

Study partner-associated CDR-SB differences increased with the proportion of comparator-arm participants reassigned at follow-up to the female living-with profile. In the AD-primary cohort, differences ranged from 0.014 points at 10% reassignment to 0.071 points at 50% reassignment, with similar but less precise patterns in AAT-relevant cohorts (Figure 2, Table 2A). Achieved target-profile increases are reported in Table S2A.

**Figure 2:**
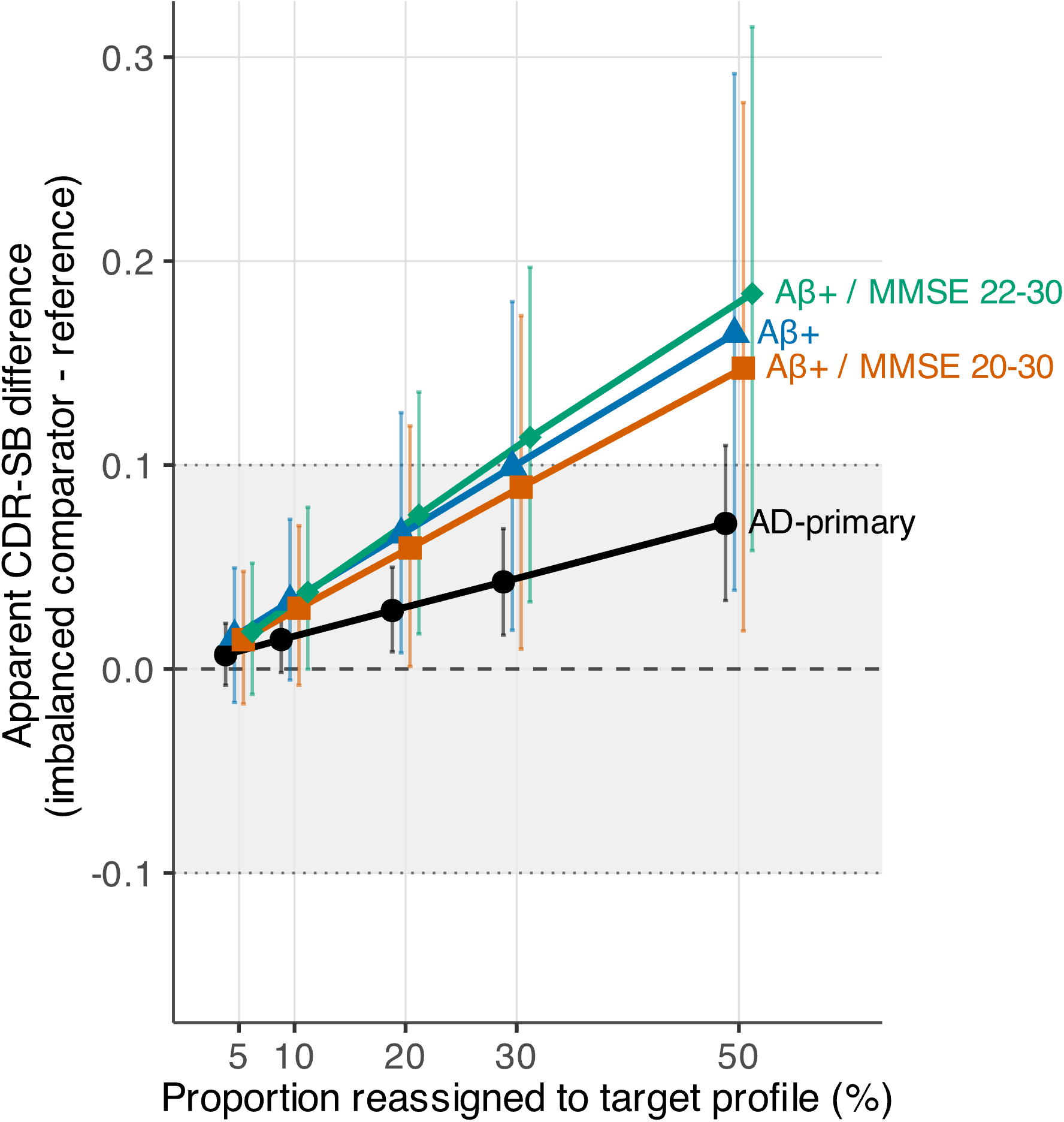
Study partner-associated CDR-SB difference under simulated follow-up imbalance in the female living-with profile. Mean study partner-associated CDR-SB difference (95% simulation interval) between the imbalanced comparator arm and the reference arm, as a function of the proportion of comparator-arm participants reassigned at follow-up to the female living-with target profile. The relationship category at follow-up is left unchanged in both arms; only study partner sex and lives-with contact are reassigned in the imbalanced comparator arm. The shaded band indicates the ±0.10 CDR-SB range, corresponding to the smallest tipping-point threshold evaluated, for visual reference. The female living-with profile was observed in ∼47–50% of participants across cohorts, making arm-level imbalance in this profile plausible in observational settings. The study partner-associated difference is expressed as imbalanced comparator arm minus reference arm; reassignment in the reference arm rather than the comparator arm would reverse the sign without changing the magnitude. **Abbreviations:** CDR-SB, Clinical Dementia Rating Sum of Boxes; Aβ+, amyloid-positive; MMSE-eq, MMSE-equivalent eligibility score range; SI, simulation interval.

### Tipping-point analysis for the female living-with profile

In the AD-primary cohort, the median net profile imbalance required to generate a 0.10-point study partner-associated CDR-SB difference was 22.5% (95% SI 15.4 to 41.1%). For a 0.20-point difference, the required imbalance was 45.0% (95% SI 30.8 to 82.3%). For a 0.30-point difference, 67.4% (95% SI 46.3 to 123.4%) was required (Figure 3). Under the primary calibration model, generating a 0.45-point study partner-associated difference, equivalent to the lecanemab treatment effect in the Clarity AD trial, required a net imbalance of 101.1% (95% SI 69.4 to 185.1%), exceeding the theoretical maximum and achievable in fewer than half of simulation iterations (proportion requiring ≤100% imbalance: 0.48) (Table 2B).

**Figure 3:**
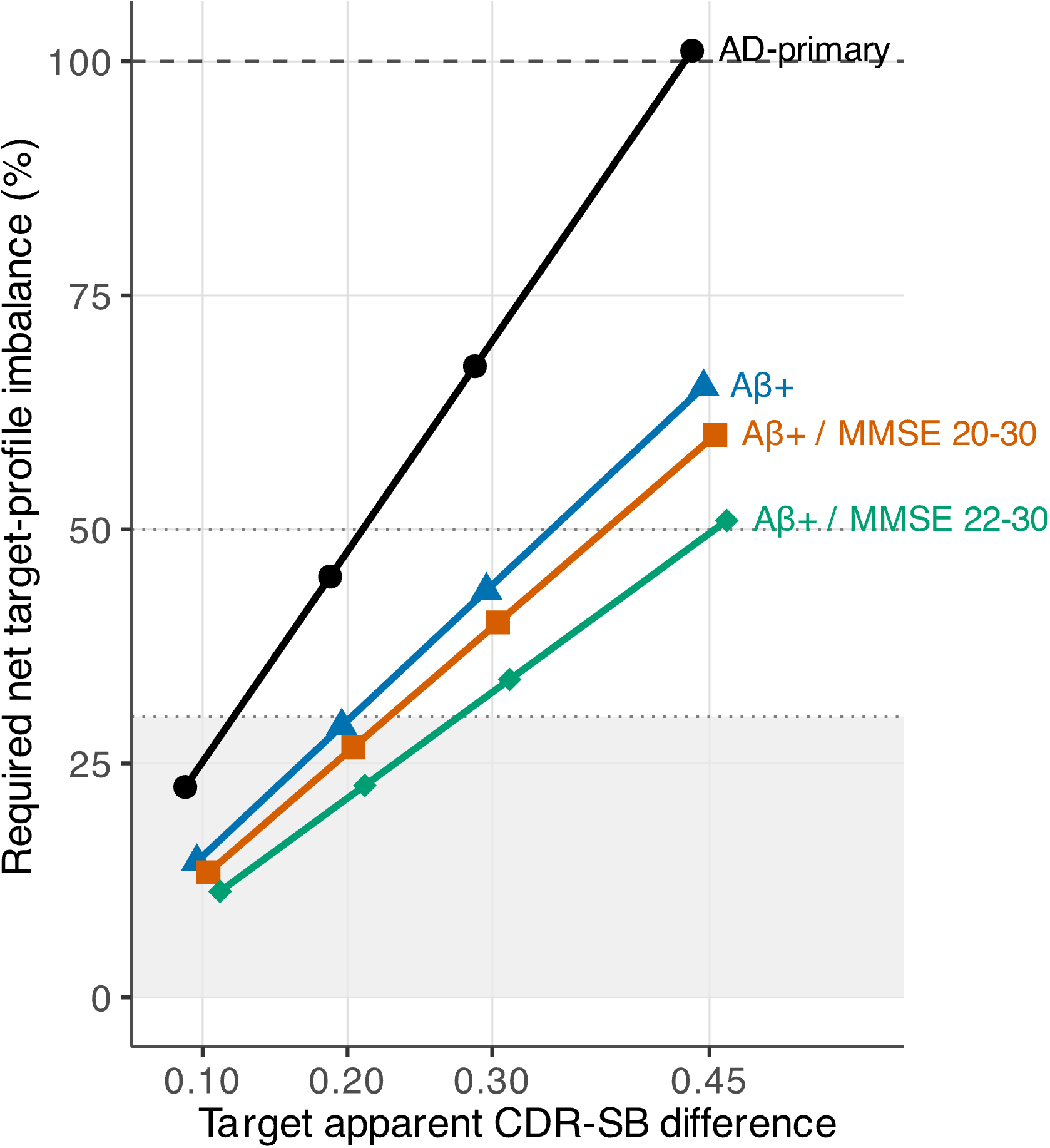
Tipping-point analysis: net female living-with profile imbalance required to generate prespecified study partner-associated CDR-SB differences. Median required net target-profile imbalance needed to generate study partner-associated CDR-SB differences of 0.10, 0.20, 0.30, and 0.45 points under the female living-with contrast, shown separately for each analytic cohort. The 0.45-point threshold corresponds to the reported 18-month CDR-SB group difference in Clarity AD. Values exceeding 100% are theoretically unachievable. Simulation intervals are reported in Table 2B. **Abbreviations:** CDR-SB, Clinical Dementia Rating Sum of Boxes; Aβ+, amyloid-positive; MMSE-eq, MMSE-equivalent eligibility score range; SI, simulation interv

In the amyloid-positive and trial-like cohorts, required imbalances were lower in point estimate but with substantially wider simulation intervals reflecting coefficient uncertainty in these smaller subsets rather than pseudo-arm sampling variability (Table 2B).

### Maximal stress-test scenarios

In the female adult-child living-with stress-test scenario, study partner-associated CDR-SB differences were larger than those observed for the common female living-with profile in the AD-primary cohort but remained modest at plausible reassignment levels. Even 50% reassignment generated 0.131 points (95% SI 0.080 to 0.183). Because the adult-child coefficient was positive in the AD-primary cohort but negative and imprecise in the amyloid-positive subsets, this profile should be interpreted as a prespecified stress-test contrast rather than as a uniformly high-score-direction or worst-case profile across all cohorts. Full results are presented in Table S2B and Table S2C.

### Decomposition of simulation uncertainty

Decomposition analyses confirmed that wide simulation intervals in the AAT-relevant cohorts were driven primarily by coefficient uncertainty rather than pseudo-arm sampling variability; full results are provided in the Table S2D.

## Discussion

This study used NACC data, calibration modeling, and Monte Carlo simulation to quantify study partner-associated components of 18-month CDR-SB change. Three findings emerge. First, naturally occurring profile changes generated negligible cohort-level components. Second, simulated follow-up imbalance in the common female living-with profile generated small, bounded CDR-SB differences in the AD-primary cohort. Third, under the primary calibration model, generating a Clarity AD-sized study partner-associated difference through this mechanism required net profile imbalance exceeding 100% in the AD-primary cohort. These results suggest that cross-sectional study partner associations translated into limited influence on 18-month CDR-SB change under the primary AD-primary model and observed profile-change patterns, while supporting study partner profile collection as a low-burden sensitivity variable.

Our calibration coefficients in the AD-primary cohort were directionally consistent with those reported by the prior NACC study (8), though generally smaller in magnitude. In that study, a female informant was associated with a CDR-SB score 0.20 points higher, an adult child informant with a score 0.39 points higher, and cohabitation with a score 0.57 points higher. In our primary cohort, estimates for the main target terms were directionally consistent but generally smaller, likely reflecting differences in cohort composition and covariate adjustment strategy. Importantly, the direction of all main effects was preserved, and our expanded calibration model yielded coefficients closer to those of the prior NACC study (8), with the lives-with coefficient increasing to 0.403 points. This indicates that tipping-point estimates based on the primary model should be interpreted as model-specific rather than as fixed thresholds.

Sensitivity analyses reinforced that the simulation estimates are calibration-association sensitivity estimates, not causal effects of changing study partners. Because the reassignment and tipping-point scenarios evaluate between-arm differences in follow-up profile distributions, level calibration coefficients are the relevant estimands for these simulations. Participant fixed-effect and change-on-change models address a different within-person estimand and showed less stable contact-frequency and lives-with estimates, consistent with selective within-person profile changes and residual confounding by care needs or disease severity.

Prior work has examined informant replacement within clinical trials (7) and between-person differences in study partner type (9). The present analyses address a separate question: whether within-person changes in broader study partner profile attributes, such as sex, relationship, and contact frequency, generate measurable study partner-associated components of 18-month CDR-SB change. In this dataset, the relevant transitions, including spouse-to-adult-child transitions, were too rare to do so under natural conditions.

In real-world AAT studies, CDR-SB is a central outcome measure (4), and differential baseline-to-follow-up changes in study partner profiles may contribute to between-group CDR-SB differences that are not explained by clinical progression alone. Static profile prevalence differences should largely cancel out in change-based analyses if profiles remain stable. Our results suggest that this concern is measurable but bounded when follow-up imbalance involves the common female living-with profile: even 30% to 50% reassignment, corresponding to a large between-arm prevalence difference, generated study partner-associated differences of approximately 0.04 to 0.07 points. These values are small relative to the reported Clarity AD group difference but may be relevant in subgroup, sensitivity, or health-economic analyses (15).

The amyloid-positive and trial-like cohort estimates should be regarded as exploratory because of smaller calibration samples and fewer baseline-to-18-month pairs. Median tipping points were lower than in the AD-primary cohort, but simulation intervals were wide because of coefficient imprecision, and adult-child coefficients were negative and imprecise in amyloid-positive subsets. Thus, current NACC data neither establish nor exclude treatment-sized study partner-associated differences in biomarker-confirmed real-world populations, a relevant uncertainty for AAT implementation studies (16). Systematic collection of study partner profile variables and sensitivity analyses are therefore particularly relevant for external comparators and target trial emulations.

Several limitations warrant consideration. First, calibration coefficients are associations, not causal effects of changing a study partner, and may include confounding by severity, care needs, living situation, and family structure. Contact frequency is particularly difficult to interpret causally because greater impairment may increase cohabitation or contact frequency. Second, the tipping-point and reassignment scenarios isolate study partner profile shifts and assume no concurrent arm-level differences in clinical severity, age, comorbidity, or other prognostic factors; we therefore interpret them as sensitivity contrasts, and not as direct corrections. Third, selective amyloid biomarker availability limits generalizability of AAT-relevant cohort coefficients. Fourth, the zCog covariate assumes that standardized MMSE and MoCA values capture comparable cognitive severity, although subcohort-specific cognitive scaling was examined in sensitivity analysis. Fifth, the analytic dataset did not include harmonized direct measures of income, employment, insurance coverage, or financial security across all included UDS versions, so residual socioeconomic confounding cannot be excluded (17). Sixth, external datasets may lack comparable study partner variables; the simulation bounds reported here are sensitivity estimates and should not be applied as direct corrections. Finally, the main and stress-test profile contrasts should be interpreted relative to profile prevalence and coefficient stability: the female living-with profile was common, whereas the female adult-child living-with profile was rare and should be viewed as a prespecified stress-test contrast.

Overall, study partner profile changes appear unlikely to materially affect 18-month CDR-SB interpretation in the large AD-primary cohort under observed or plausible profile-change scenarios, but uncertainty in biomarker-confirmed cohorts supports systematic study partner profile collection and sensitivity analyses in observational and external-comparator CDR-SB studies for anti-amyloid therapy evaluation.

## Acknowledgements

The NACC database is funded by NIA/NIH Grant U24 AG072122. NACC data are contributed by the NIA-funded ADRCs: P30 AG062429 (PI James Brewer, MD, PhD), P30 AG066468 (PI Oscar Lopez, MD), P30 AG062421 (PI Bradley Hyman, MD, PhD), P30 AG066509 (PI Thomas Grabowski, MD), P30 AG066514 (PI Mary Sano, PhD), P30 AG066530 (PI Helena Chui, MD), P30 AG066507 (PI Marilyn Albert, PhD), P30 AG066444 (PI David Holtzman, MD), P30 AG066518 (PI Lisa Silbert, MD, MCR), P30 AG066512 (PI Thomas Wisniewski, MD), P30 AG066462 (PI Scott Small, MD), P30 AG072979 (PI David Wolk, MD), P30 AG072972 (PI Charles DeCarli, MD), P30 AG072976 (PI Andrew Saykin, PsyD), P30 AG072975 (PI Julie A. Schneider, MD, MS), P30 AG072978 (PI Ann McKee, MD), P30 AG072977 (PI Robert Vassar, PhD), P30 AG066519 (PI Frank LaFerla, PhD), P30 AG062677 (PI Ronald Petersen, MD, PhD), P30 AG079280 (PI Jessica Langbaum, PhD), P30 AG062422 (PI Gil Rabinovici, MD), P30 AG066511 (PI Allan Levey, MD, PhD), P30 AG072946 (PI Linda Van Eldik, PhD), P30 AG062715 (PI Sanjay Asthana, MD, FRCP), P30 AG072973 (PI Russell Swerdlow, MD), P30 AG066506 (PI Glenn Smith, PhD, ABPP), P30 AG066508 (PI Stephen Strittmatter, MD, PhD), P30 AG066515 (PI Victor Henderson, MD, MS), P30 AG072947 (PI Suzanne Craft, PhD), P30 AG072931 (PI Henry Paulson, MD, PhD), P30 AG066546 (PI Sudha Seshadri, MD), P30 AG086401 (PI Erik Roberson, MD, PhD), P30 AG086404 (PI Gary Rosenberg, MD), P20 AG068082 (PI Angela Jefferson, PhD), P30 AG072958 (PI Heather Whitson, MD), P30 AG072959 (PI James Leverenz, MD).

Some authors’ affiliation (KS, YN, TI), “*Dementia Inclusion and Therapeutics*,” is an endowed department at the University of Tokyo Hospital funded by Effissimo Capital Management Pte Ltd.

AI-assisted tools were used for language editing; authors take full responsibility for the content.

## Consent Statement

Informed consent is not required for this type of study.

## Data Availability

NACC UDS data are available to qualified investigators through the National Alzheimer’s Coordinating Center after submission and approval of a research proposal and execution of a data use agreement.

