## Supplementary Materials for "Study partner profile effects on CDR-SB change in anti-amyloid therapy evaluation"

**Supplementary Methods**

Supplementary Methods include details of cognitive severity construction, amyloid status coding, contact frequency coding, model fitting procedures, Monte Carlo simulation procedures, and sensitivity analyses.

*Cognitive severity covariate*

A composite cognitive severity variable, zCog, was constructed from raw MMSE and MoCA scores standardized within the AD-primary cohort. When both were available, the MMSE z-score was used; when missing, MoCA was substituted. A separate sensitivity analysis re-standardized MMSE and MoCA within each AAT-relevant subcohort to assess whether subcohort-specific cognitive scaling materially affected study partner-associated coefficients. An MMSE-equivalent eligibility score, which may incorporate MoCA-to-MMSE conversion derived in the preprocessing step, was used exclusively for defining trial-like AAT eligibility cuts. It was not used as the calibration covariate.

*Amyloid status*

Amyloid status was incorporated as a covariate in the expanded calibration model and used to define the amyloid-positive cohort. Amyloid status was coded as a three-level variable: positive (at least one positive PET or CSF result), non-positive available (both available results negative), and unknown (no biomarker available). Participants with unknown amyloid status were not collapsed with the non-positive category.

*Contact frequency coding:*

Direct cohabitation information was always prioritized and classified as lives-with. When a derived contact frequency variable was available, it was used as the primary source; when it was missing, an in-person visit frequency variable was used as a fallback. This hierarchical rule was implemented to prevent the fallback variable from overriding non-missing primary source values, as the two variables use overlapping but not identical coding schemes across UDS versions.

*Calibration model convergence*

When the full model failed to converge, a prespecified sequence of fallback models was attempted: center as fixed effect instead of random, then removal of the random slope, then removal of the center term entirely. A sensitivity analysis additionally fitted an expanded calibration model including age, education, race and ethnicity, primary language, APOE ε4 carrier status, and amyloid status as additional fixed-effect covariates. **The random intercept and random slope for time were specified as uncorrelated to improve model stability, particularly in the smaller AAT-relevant cohorts where estimating a free correlation parameter may lead to convergence issues.** Convergence warnings for singular fits were allowed given the large number of random effects relative to some subcohort sizes.

*Monte Carlo simulation*

The matrix V was verified to be positive definite before use in simulation. To separate the contribution of coefficient uncertainty from pseudo-arm sampling variability, the between-arm simulation was repeated under two conditions: beta drawn, in which a new coefficient vector was sampled at each iteration from the multivariate normal distribution, and beta fixed, in which coefficients were held at their point estimates across all iterations. The simulation was additionally repeated for pseudo-arm sizes of 250, 500, 700, and 900 participants per arm, using 2,000 iterations per condition.

The study partner-associated coefficient vector β and its variance-covariance matrix V were extracted from the calibration model. The study partner-associated terms comprised: informant sex, adult child relationship (versus spouse or partner), other relationship (versus spouse or partner), weekly-or-more contact (versus less than weekly), daily contact (versus less than weekly), lives-with cohabitation (versus less than weekly), and the two participant-sex-by-relationship interaction terms.

Design vector construction: For each participant with a complete baseline-to-follow-up pair, the study partner profile design vectors at baseline (X_baseline) and follow-up (X_followup) were constructed using the model matrix corresponding to the study partner profile formula. The difference vector ΔX = X_followup - X_baseline captures the change in study partner profile between the two timepoints.

Simulation procedure: At each Monte Carlo iteration, a coefficient vector β(m) was drawn from a multivariate normal distribution with mean β and covariance matrix V. The study partner-associated component for each participant was then computed as: *component_i(m) = ΔX_i · β(m)*. The cohort-level mean component was computed across participants at each iteration. Summary statistics were computed across iterations, including the mean, median, standard deviation, and 2.5th and 97.5th percentiles of the distribution. The proportion of iterations in which the absolute value of the mean component exceeded thresholds of 0.10, 0.20, 0.30, and 0.45 CDR-SB points was also reported. The primary analyses used 10,000 Monte Carlo iterations.

*Coefficient interpretation*

The coefficients from the calibration model are calibration associations, not causal effects of changing a study partner. They may include confounding by patient severity and care needs, particularly for contact frequency, because patients with greater impairment may be more likely to live with or have frequent contact with their study partner. Results are therefore reported as study partner-associated CDR-SB components rather than as measurement bias estimates or causal switching effects. Additional simulation scenarios were evaluated as described below.

*Sensitivity analyses:*
The following prespecified sensitivity analyses were conducted. An expanded calibration model added age, education, race and ethnicity, primary language, *APOE* ε4 status, cognition source, and amyloid status as additional covariates. An alternative follow-up window of 9 to 27 months was evaluated. A quadratic zCog term was added to assess non-linearity of the cognitive severity adjustment. Visits with informant reliability concerns were excluded when the relevant variable was available. A participant fixed-effect model was fitted using ordinary least squares with participant indicators, including informant sex, relationship, and contact as fixed effects but omitting the participant sex by relationship interaction, because within-person sex-specific relationship transitions are sparse and produce unstable estimates. A change-on-change model regressed the change in CDR-SB on changes in study partner profile variables between baseline and follow-up. An external coefficient sensitivity analysis used published estimates from Vargas-Gonzalez et al. (2024), sampled independently from normal distributions because the published covariance matrix was not available.

**Table S1: Calibration model and sensitivity analysis coefficients for the association between study partner profile characteristics and CDR-SB levels**

(A) Main calibration model: study partner-associated coefficients by cohort

| **Cohort** | **Term** | **Standard error** | **Estimate (95% Ci)** |
| --- | --- | --- | --- |
| AD-primary | Female informant | 0.044 | 0.143 (0.055 to 0.230) |
|  | Child vs spouse/partner | 0.071 | 0.165 (0.025 to 0.305) |
|  | Other vs spouse/partner | 0.084 | -0.295 (-0.460 to -0.131) |
|  | Weekly-or-more vs <weekly | 0.085 | 0.196 (0.029 to 0.363) |
|  | Daily vs <weekly | 0.088 | 0.206 (0.033 to 0.379) |
|  | Lives with vs <weekly | 0.092 | 0.303 (0.122 to 0.483) |
|  | Patient female x child | 0.087 | 0.087 (-0.085 to 0.258) |
|  | Patient female x other | 0.104 | 0.146 (-0.058 to 0.351) |
| Aβ+ | Female informant | 0.153 | 0.316 (0.016 to 0.616) |
|  | Child vs spouse/partner | 0.271 | -0.070 (-0.602 to 0.461) |
|  | Other vs spouse/partner | 0.309 | -0.479 (-1.085 to 0.127) |
|  | Weekly-or-more vs <weekly | 0.338 | 0.410 (-0.253 to 1.073) |
|  | Daily vs <weekly | 0.354 | -0.205 (-0.900 to 0.489) |
|  | Lives with vs <weekly | 0.367 | 0.378 (-0.341 to 1.096) |
|  | Patient female x child | 0.314 | 0.181 (-0.435 to 0.796) |
|  | Patient female x other | 0.377 | 0.291 (-0.448 to 1.030) |
| Aβ+ / MMSE-20-30 | Female informant | 0.154 | 0.287 (-0.015 to 0.590) |
|  | Child vs spouse/partner | 0.281 | -0.228 (-0.779 to 0.324) |
|  | Other vs spouse/partner | 0.320 | -0.628 (-1.255 to -0.000) |
|  | Weekly-or-more vs <weekly | 0.328 | 0.498 (-0.146 to 1.141) |
|  | Daily vs <weekly | 0.348 | -0.019 (-0.700 to 0.663) |
|  | Lives with vs <weekly | 0.367 | 0.464 (-0.256 to 1.185) |
|  | Patient female x child | 0.319 | 0.350 (-0.275 to 0.976) |
|  | Patient female x other | 0.383 | 0.384 (-0.365 to 1.134) |
| Aβ+ / MMSE 22-30 | Female informant | 0.158 | 0.364 (0.053 to 0.675) |
|  | Child vs spouse/partner | 0.285 | -0.159 (-0.719 to 0.400) |
|  | Other vs spouse/partner | 0.325 | -0.557 (-1.195 to 0.081) |
|  | Weekly-or-more vs <weekly | 0.329 | 0.456 (-0.190 to 1.101) |
|  | Daily vs <weekly | 0.351 | 0.001 (-0.688 to 0.690) |
|  | Lives with vs <weekly | 0.371 | 0.520 (-0.207 to 1.248) |
|  | Patient female x child | 0.327 | 0.228 (-0.413 to 0.870) |
|  | Patient female x other | 0.392 | 0.242 (-0.526 to 1.010) |

(B) Sensitivity analyses: study partner-associated coefficients and observed-transition components, AD-primary cohort

| **Analysis** | **Term** | **Standard error** | **Estimate (95% Ci)** |
| --- | --- | --- | --- |
| Expanded calibration model | Female informant | 0.048 | 0.170 (0.075 to 0.264) |
|  | Child vs spouse/partner | 0.077 | 0.214 (0.062 to 0.366) |
|  | Other vs spouse/partner | 0.091 | -0.209 (-0.388 to -0.031) |
|  | Weekly-or-more vs <weekly | 0.091 | 0.232 (0.053 to 0.411) |
|  | Daily vs <weekly | 0.095 | 0.305 (0.119 to 0.492) |
|  | Lives with vs <weekly | 0.099 | 0.403 (0.208 to 0.598) |
|  | Patient female x child | 0.094 | 0.018 (-0.167 to 0.203) |
|  | Patient female x other | 0.113 | 0.146 (-0.076 to 0.368) |
| Quadratic cognitive severity adjustment | Female informant | 0.044 | 0.145 (0.057 to 0.232) |
|  | Child vs spouse/partner | 0.072 | 0.182 (0.041 to 0.322) |
|  | Other vs spouse/partner | 0.084 | -0.295 (-0.461 to -0.130) |
|  | Weekly-or-more vs <weekly | 0.084 | 0.196 (0.031 to 0.362) |
|  | Daily vs <weekly | 0.088 | 0.228 (0.056 to 0.400) |
|  | Lives with vs <weekly | 0.092 | 0.320 (0.141 to 0.500) |
|  | Patient female x child | 0.088 | 0.090 (-0.083 to 0.263) |
|  | Patient female x other | 0.105 | 0.155 (-0.051 to 0.361) |
| Excluding unreliable informant visits | Female informant | 0.045 | 0.141 (0.052 to 0.230) |
|  | Child vs spouse/partner | 0.072 | 0.161 (0.019 to 0.303) |
|  | Other vs spouse/partner | 0.086 | -0.302 (-0.471 to -0.134) |
|  | Weekly-or-more vs <weekly | 0.088 | 0.193 (0.020 to 0.366) |
|  | Daily vs <weekly | 0.091 | 0.195 (0.016 to 0.374) |
|  | Lives with vs <weekly | 0.095 | 0.300 (0.113 to 0.487) |
|  | Patient female x child | 0.088 | 0.099 (-0.075 to 0.272) |
|  | Patient female x other | 0.107 | 0.156 (-0.053 to 0.366) |
| Participant fixed-effect model | Female informant | 0.074 | 0.284 (0.140 to 0.429) |
|  | Child vs spouse/partner | 0.095 | -0.128 (-0.315 to 0.059) |
|  | Other vs spouse/partner | 0.112 | -0.210 (-0.428 to 0.009) |
|  | Weekly-or-more vs <weekly | 0.123 | 0.020 (-0.221 to 0.260) |
|  | Daily vs <weekly | 0.131 | -0.027 (-0.284 to 0.230) |
|  | Lives with vs <weekly | 0.139 | -0.233 (-0.505 to 0.040) |
| Change-on-change model | Delta female informant | 0.122 | 0.099 (-0.140 to 0.337) |
|  | Delta child relationship | 0.162 | -0.253 (-0.571 to 0.065) |
|  | Delta other relationship | 0.192 | -0.193 (-0.569 to 0.183) |
|  | Delta weekly-or-more contact | 0.218 | -0.024 (-0.452 to 0.404) |
|  | Delta daily contact | 0.229 | -0.143 (-0.592 to 0.307) |
|  | Delta lives-with contact | 0.245 | -0.455 (-0.935 to 0.026) |
| Alternative follow-up window (9-27 months) | Observed transition mean component |  | 0.001 (0.001 to 0.002) |
| External coefficients (Vargas-Gonzalez et al., 2024) | Observed transition mean component |  | 0.002 (-0.000 to 0.004) |

**(A)** Point estimates, standard errors, and 95% confidence intervals for study partner-associated fixed-effect terms from the primary linear mixed-effects calibration model, fitted separately for each analytic cohort. Confidence intervals were derived from the model-based standard errors. Terms whose confidence intervals do not include zero are statistically distinguishable from zero at the 5% level.

**(B)** Point estimates, standard errors, and 95% confidence intervals for study partner-associated terms across prespecified sensitivity analyses, fitted in the AD-primary cohort unless otherwise noted. For the alternative follow-up window and external coefficient sensitivity analyses, the metric reported is the cohort-level mean study partner-associated CDR-SB component under observed transitions rather than individual model coefficients. Standard errors are not available for the external coefficient sensitivity analysis because published covariance information was not available and coefficients were sampled independently.

**Abbreviations:** CDR-SB, Clinical Dementia Rating Sum of Boxes; Aβ+, amyloid-positive; MMSE-eq, MMSE-equivalent eligibility score range; CI, confidence interval; SE, standard error; AD, Alzheimer's disease.

**Table S2: Supplementary simulation results: achieved profile prevalence, maximal stress-test between-arm imbalance, maximal stress-test tipping-point analysis, decomposition of simulation uncertainty, and contact source diagnostic.**

(A). Achieved target-profile prevalence under simulated between-arm imbalance in the female living-with profile

| **Cohort** | **Requested reassignment** | **Final female living-with prevalence (%)** | **Actual target-profile increase (percentage points)** |
| --- | --- | --- | --- |
| AD-primary | 5% | 51.7 | 4.8 |
|  | 10% | 56.9 | 10.0 |
|  | 20% | 66.9 | 20.0 |
|  | 30% | 76.8 | 30.0 |
|  | 50% | 96.6 | 49.7 |
| Aβ+ | 5% | 52.7 | 4.8 |
|  | 10% | 58.0 | 10.0 |
|  | 20% | 67.9 | 20.0 |
|  | 30% | 78.0 | 30.0 |
|  | 50% | 97.4 | 49.5 |
| Aβ+ / MMSE 20-30 | 5% | 54.0 | 4.8 |
|  | 10% | 59.1 | 10.0 |
|  | 20% | 69.2 | 20.0 |
|  | 30% | 79.1 | 30.0 |
|  | 50% | 98.3 | 49.1 |
| Aβ+ / MMSE 22-30 | 5% | 54.4 | 4.8 |
|  | 10% | 59.6 | 10.0 |
|  | 20% | 69.6 | 20.0 |
|  | 30% | 79.6 | 30.0 |
|  | 50% | 98.5 | 48.9 |

Values represent the mean final prevalence of the female living-with profile in the imbalanced comparator arm after reassignment, averaged across Monte Carlo iterations. The actual target-profile increase column reports the achieved increase in percentage points relative to the reference arm. Requested reassignment refers to the proportion of comparator-arm participants reassigned to the female living-with profile.

(B). Between-arm imbalance results: maximal female adult-child living-with profile stress-test scenario

| **Cohort** | **Requested reassignment** | **Mean study partner-associated CDR-SB difference (95% SI)** | **Final target-profile prevalence (%)** |
| --- | --- | --- | --- |
| AD-primary | 5% | 0.013 (-0.003 to 0.029) | 9.0 |
|  | 10% | 0.026 (0.009 to 0.045) | 14.2 |
|  | 20% | 0.053 (0.028 to 0.079) | 24.2 |
|  | 30% | 0.079 (0.046 to 0.113) | 34.2 |
|  | 50% | 0.131 (0.080 to 0.183) | 54.2 |
| Aβ+ | 5% | 0.010 (-0.026 to 0.047) | 7.1 |
|  | 10% | 0.021 (-0.028 to 0.070) | 12.3 |
|  | 20% | 0.043 (-0.039 to 0.126) | 22.3 |
|  | 30% | 0.064 (-0.054 to 0.182) | 32.4 |
|  | 50% | 0.105 (-0.085 to 0.299) | 52.3 |
| Aβ+ / MMSE 20-30 | 5% | 0.006 (-0.030 to 0.044) | 7.0 |
|  | 10% | 0.012 (-0.041 to 0.063) | 12.2 |
|  | 20% | 0.025 (-0.063 to 0.112) | 22.2 |
|  | 30% | 0.037 (-0.089 to 0.162) | 32.2 |
|  | 50% | 0.062 (-0.146 to 0.264) | 52.2 |
| Aβ+ / MMSE 22-30 | 5% | 0.009 (-0.028 to 0.046) | 7.1 |
|  | 10% | 0.018 (-0.034 to 0.070) | 12.3 |
|  | 20% | 0.035 (-0.054 to 0.125) | 22.3 |
|  | 30% | 0.053 (-0.077 to 0.181) | 32.3 |
|  | 50% | 0.088 (-0.123 to 0.298) | 52.3 |

Values represent the mean study partner-associated CDR-SB difference (95% simulation interval) between the imbalanced comparator arm and the reference arm when the specified proportion of comparator-arm participants is reassigned to the female adult-child living-with target profile. This profile was observed in approximately 4% of participants in the AD-primary cohort and should be interpreted as an upper-bound stress test rather than a plausible observational imbalance. The final target-profile prevalence column reports the absolute prevalence of the female adult-child living-with profile in the comparator arm after reassignment.

(C). Tipping-point analysis: maximal female adult-child living-with profile stress-test scenario

| **Cohort** | **Target study partner-associated CDR-SB difference (points)** | **Required net imbalance, median (95% SI)** | **Proportion of iterations requiring <=100% imbalance** |
| --- | --- | --- | --- |
| AD-primary | 0.10 | 15.2% (11.1 to 24.2%) | 1.000 |
|  | 0.20 | 30.5% (22.2 to 48.4%) | 1.000 |
|  | 0.30 | 45.7% (33.4 to 72.6%) | 0.998 |
|  | 0.45 | 68.5% (50.1 to 108.9%) | 0.952 |
| Aβ+ | 0.10 | 14.1% (6.1 to 221.3%) | 0.943 |
|  | 0.20 | 28.1% (12.1 to 442.6%) | 0.883 |
|  | 0.30 | 42.2% (18.2 to 663.8%) | 0.819 |
|  | 0.45 | 63.2% (27.3 to 995.7%) | 0.711 |
| Aβ+ / MMSE 20-30 | 0.10 | 14.3% (6.0 to 277.6%) | 0.937 |
|  | 0.20 | 28.6% (12.0 to 555.3%) | 0.876 |
|  | 0.30 | 42.9% (18.0 to 832.9%) | 0.809 |
|  | 0.45 | 64.3% (26.9 to 1249.4%) | 0.693 |
| Aβ+ / MMSE 22-30 | 0.10 | 12.0% (5.5 to 164.4%) | 0.959 |
|  | 0.20 | 24.0% (10.9 to 328.8%) | 0.915 |
|  | 0.30 | 36.1% (16.4 to 493.2%) | 0.863 |
|  | 0.45 | 54.1% (24.6 to 739.8%) | 0.784 |

Values represent the median required net target-profile imbalance (95% simulation interval) across Monte Carlo iterations needed to generate the specified study partner-associated CDR-SB difference under the female adult-child living-with profile contrast. The proportion of iterations requiring 100% or less imbalance indicates the feasibility of the specified difference under this contrast. Values exceeding 100% are theoretically unachievable. Because the female adult-child living-with profile was observed in approximately 4% of participants, these estimates represent an upper-bound stress test.

(D). Decomposition of simulation uncertainty: 95% simulation interval width by beta-sampling condition and pseudo-arm size.

| **Cohort** | **Requested reassignment** | **Beta-sampling condition** | **Pseudo-arm size (n)** | **95% simulation interval width** | **Mean study partner-associated CDR-SB difference** |
| --- | --- | --- | --- | --- | --- |
| AD-primary | 30% | Drawn | 250 | 0.051 | 0.043 |
|  | 30% | Fixed | 250 | 0.029 | 0.043 |
|  | 30% | Drawn | 900 | 0.044 | 0.043 |
|  | 30% | Fixed | 900 | 0.015 | 0.043 |
|  | 50% | Drawn | 250 | 0.077 | 0.071 |
|  | 50% | Fixed | 250 | 0.028 | 0.071 |
|  | 50% | Drawn | 900 | 0.070 | 0.072 |
|  | 50% | Fixed | 900 | 0.016 | 0.072 |
| Aβ+ | 30% | Drawn | 250 | 0.157 | 0.098 |
|  | 30% | Fixed | 250 | 0.054 | 0.099 |
|  | 30% | Drawn | 900 | 0.149 | 0.099 |
|  | 30% | Fixed | 900 | 0.029 | 0.099 |
|  | 50% | Drawn | 250 | 0.255 | 0.164 |
|  | 50% | Fixed | 250 | 0.055 | 0.164 |
|  | 50% | Drawn | 900 | 0.250 | 0.165 |
|  | 50% | Fixed | 900 | 0.029 | 0.165 |
| Aβ+ / MMSE 20-30 | 30% | Drawn | 250 | 0.165 | 0.090 |
|  | 30% | Fixed | 250 | 0.052 | 0.088 |
|  | 30% | Drawn | 900 | 0.151 | 0.090 |
|  | 30% | Fixed | 900 | 0.028 | 0.089 |
|  | 50% | Drawn | 250 | 0.259 | 0.146 |
|  | 50% | Fixed | 250 | 0.052 | 0.146 |
|  | 50% | Drawn | 900 | 0.254 | 0.149 |
|  | 50% | Fixed | 900 | 0.026 | 0.147 |
| Aβ+ / MMSE 22-30 | 30% | Drawn | 250 | 0.166 | 0.114 |
|  | 30% | Fixed | 250 | 0.053 | 0.114 |
|  | 30% | Drawn | 900 | 0.154 | 0.115 |
|  | 30% | Fixed | 900 | 0.028 | 0.114 |
|  | 50% | Drawn | 250 | 0.254 | 0.187 |
|  | 50% | Fixed | 250 | 0.055 | 0.185 |
|  | 50% | Drawn | 900 | 0.253 | 0.186 |
|  | 50% | Fixed | 900 | 0.029 | 0.187 |

The 95% simulation interval width is defined as the difference between the 97.5th and 2.5th percentiles of the distribution of mean study partner-associated CDR-SB differences across Monte Carlo iterations. Under the beta-drawn condition, a new coefficient vector was sampled at each iteration from the multivariate normal distribution, incorporating both coefficient uncertainty and pseudo-arm sampling variability. Under the beta-fixed condition, coefficients were held at their point estimates, isolating pseudo-arm sampling variability alone. Results are shown for the common female living-with target profile at 30% and 50% requested reassignment levels and for pseudo-arm sizes of 250 and 900 participants per arm. The decomposition used 2,000 Monte Carlo iterations per condition.

(E). Contact source diagnostic: agreement between contact frequency source variables.

| **Cohort** | **Contact frequency source used** | **Analytic lives-with rate (%)** | **Visits where primary source indicated non-lives-with but fallback indicated lives-with, n** |
| --- | --- | --- | --- |
| AD-primary | *NACCINCNTFQ* | 69.9 | 32,653 |
| Aβ+ | *NACCINCNTFQ* | 79.1 | 2,913 |
| Aβ+ / MMSE 20-30 | *NACCINCNTFQ* | 79.1 | 2,374 |
| Aβ+ / MMSE 22-30 | *NACCINCNTFQ* | 79.1 | 2,096 |

The source-aware hierarchy operated in three steps: first, visits where INLIVWTH equal to 1 were always classified as lives-with regardless of other variables; second, for remaining visits, NACCINCNTFQ was used as the primary contact frequency source when available and non-missing, with code 8 indicating lives-with; third, when NACCINCNTFQ was missing, INVISITS was used as fallback, with INVISITS equal to 8 also indicating lives-with. The contact frequency source column reports the primary frequency source used after the INLIVWTH cohabitation override. The analytic lives-with rate in this table reflects visits where NACCINCNTFQ was the active contact frequency source and differs slightly from the overall lives-with rate reported in Table 1, which includes visits classified as lives-with via direct cohabitation information regardless of source.

**Abbreviations:** Abeta+, amyloid-positive; MMSE-eq, MMSE-equivalent eligibility score range; NACCINCNTFQ, NACC-derived contact frequency variable (primary source used in this analysis); n, number

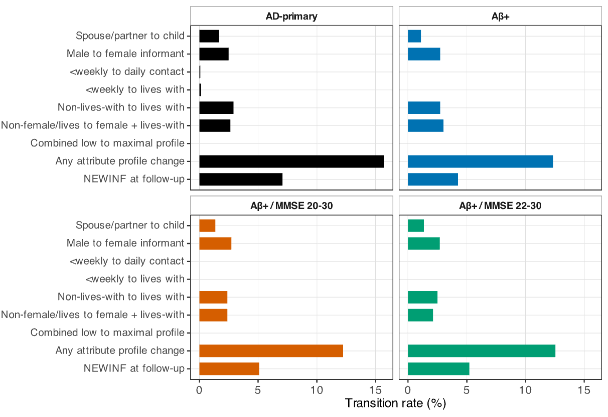
**Figure S1. Observed study partner profile transition rates between baseline and 18-month follow-up.**

Rates of specific study partner profile transitions observed between baseline and the 18-month follow-up visit, expressed as a percentage of complete baseline-to-follow-up pairs, shown separately for each analytic cohort. Transitions shown include informant replacement at follow-up, any attribute profile change, combined low-to-maximal profile transition, transition from non-female non-living-with to female living-with profile, non-lives-with to lives-with contact, less-than-weekly to lives-with contact, less-than-weekly to daily contact, male to female informant, and spouse or partner to adult child relationship. The combined low-to-maximal profile transition, defined as a simultaneous change from male informant, spouse or partner relationship, and less-than-weekly contact at baseline to female informant, adult child relationship, and daily or lives-with contact at follow-up, was not observed in any participant across all cohorts. Individual transition rates were uniformly low across all cohorts: spouse-to-child and male-to-female transitions each occurred in fewer than 3% of participants, non-lives-with-to-lives-with transitions in fewer than 3%, and transition to the female living-with profile in fewer than 3%. These low rates are consistent with the negligible cohort-level study partner-associated CDR-SB components estimated under naturally occurring profile changes.

**Abbreviations:** Aβ+, amyloid-positive; MMSE-eq, MMSE-equivalent eligibility score range; NEWINF, new informant indicator.
